# Pregnancies complicated by high maternal BMI and placental inflammation have lower placental efficiency and infant Apgar scores at preterm

**DOI:** 10.64898/2026.11.22.26368693

**Authors:** Hailey Scott, Enrrico Bloise, Kristin L Connor

## Abstract

Inflammation in pregnancy may elicit adverse pregnancy and newborn health outcomes. Obesity and chorioamnionitis are two prevalent conditions that can contribute to a pro-inflammatory environment in pregnancy. However, whether high placental inflammation identifies pregnancies complicated by maternal overweight/obesity or chorioamnionitis that are at greater risk of adverse outcomes, particularly in the absence of other major maternal comorbidities, remains unclear. Our primary objective was to examine whether high placental inflammation was associated with adverse pregnancy and neonatal outcomes in pregnancies complicated by maternal overweight/obesity or chorioamnionitis, in the absence of other major maternal comorbidities. Clinical data and placentae were obtained from 78 pregnancies across maternal prepregnancy BMI groups that delivered preterm (with and without chorioamnionitis) or term, without other major maternal comorbidities. Placental cytokine and chemokine protein expression levels were assessed by Bio-Plex immunoassay, and a pro-inflammatory summary score was derived. Placental samples with pro-inflammatory summary scores in the highest tertile were defined as having high placental inflammation, and samples with scores in the bottom two tertiles as without high inflammation. We assessed the combined effects of maternal overweight/obesity and high placental inflammation, and chorioamnionitis and high placental inflammation, on pregnancy, placental, and infant outcomes, comparing outcomes to overweight/obesity or chorioamnionitis affected pregnancies without high placental inflammation. Pregnancies complicated by maternal overweight/obesity and high placental inflammation had decreased birthweight-to-placental weight ratio (p=0.003) and lower infant Apgar scores (1-minute: p=0.03; 5-minute: p=0.01), compared to pregnancies with overweight/obesity without high placental inflammation, in preterm pregnancies only. In contrast, high placental inflammation among pregnancies with normal BMI did not associate with poorer outcomes. Chorioamnionitis cases with high placental inflammation also did not have poorer placental or infant outcomes compared to chorioamnionitis cases without high placental inflammation. Among pregnancies complicated by maternal overweight/obesity but no other major comorbidities, those with inflamed placentae may be most at risk for poor birth outcomes, particularly at preterm. Understanding placental phenotypes among sub-groups at greatest risk of adversity may allow for more targeted surveillance, early intervention, and individualised prenatal care.

## Introduction

Inflammation in pregnancy associates with adverse placental and offspring outcomes^1,2,3^, including preterm birth (PTB), altered fetal growth^4^ and brain development^5-8^. Of particular interest, sterile or infectious chorioamnionitis remains a leading contributor to PTB^19^, and can cause sepsis, fetal cerebral white matter damage, and perinatal death^20^. In addition, high maternal body mass index (BMI) and obesity^8-10^, which continue to rise globally, are also associated with an inflammatory maternal environment, and increased risk for PTB^11,12^, poor placental and infant growth^13,14^, and later health programming^15-18^.

While inflammation is a known mechanism driving adversity in pregnancy, studies investigating maternal obesity often use cohorts confounded by additional comorbidities, such as hypertension and gestational diabetes. As a result, the independent contribution of obesity-associated inflammation to placental inflammatory status and offspring outcomes remains poorly understood. Further, 20-30% of individuals with obesity do not have other major metabolic alterations^21^, indicating that this population is clinically important to understand. In addition, while maternal obesity is an established risk factor for obstetrical complications, not all pregnancies affected by high BMI experience adverse outcomes^22^. It is possible that differences in placental inflammatory states may help identify pregnancies with high BMI that are at greater risk for poor outcomes.

Adverse maternal-fetal outcomes associated with high BMI^8,14-15^ or placental inflammation^3,5^ are well established. However, little is known about whether high maternal BMI in combination with increased placental cytokine and chemokine expression, in the absence of other comorbidities, may further increase the risk of adverse neonatal outcomes. Our current understanding of placental inflammation and risk for offspring adversity is predominantly grounded in assessments of placental histopathology^3^. This includes chorioamnionitis, commonly diagnosed histologically based on neutrophil infiltration of the placenta and fetal membranes^45^, with fewer studies examining placental inflammation at the biochemical level through the assessment of cytokine and chemokine expression. Consequently, it remains unclear whether differences in placental inflammatory states are associated with adverse outcomes, and whether these associations differ according to maternal BMI or chorioamnionitis status. Thus, evaluating multiple inflammatory placental cytokine/chemokine levels in pregnancies across different maternal BMI categories, with and without chorioamnionitis, is warranted to address these gaps.

Characterising whether poorer outcomes at birth occur when high placental inflammation is present or not in pregnancies with high maternal BMI, or with chorioamnionitis, may help to further pinpoint populations that need increased obstetrical monitoring and inform novel targets for intervention. Our primary objective was to determine whether high placental inflammation, defined by a high placental pro-inflammatory cytokine and chemokine summary score, was associated with more adverse pregnancy outcomes in pregnancies affected by maternal overweight/obesity or chorioamnionitis, compared with pregnancies within the same maternal condition without high placental inflammation. We also examined whether these associations were evident in pregnancies with normal maternal BMI. We hypothesised that when both maternal overweight/obesity and high placental inflammation are present, pregnancy outcomes would be poorer than when maternal overweight/obesity exist without high placental inflammation. Similarly, we also hypothesised that high placental inflammation in pregnancies complicated by chorioamnionitis would be associated with poorer pregnancy outcomes than chorioamnionitis with a lower inflammatory phenotype.

## Methods

### Study population

In this cross-sectional study, participants were recruited from Mount Sinai Hospital (MSH), Toronto (MSH-REB:17-0186-E; Carleton University-REB:106932). Maternal and pregnancy characteristics of the full cohort (n=96) have been previously described^25^. Placentae were assessed from pregnancies across maternal BMI groups and gestational ages as follows: preterm with chorioamnionitis: underweight (UW; n=3), normal weight (NW; n=8), overweight (OW; n=10), and obese (OB; n=3); preterm without chorioamnionitis: UW (n=5), NW (n=7), OW (n=7), OB (n=8); and term: UW (n=8), NW (n=6), OW (n=6), OB (n=7). Maternal prepregnancy BMI was categorised based on World Health Organization guidelines^26,27^, with exception that prepregnancy BMI<19 was considered underweight, and BMI of 19-24.9 normal weight, due to a lower number of underweight cases in the study region. Exclusion criteria included other major maternal comorbidities known to alter development and inflammatory status^25^.

### Placental inflammation

#### Placental cytokine/chemokine quantification

Protein was extracted from frozen placentae using RIPA lysis buffer with Halt Protease and Phosphatase Inhibitor Cocktail (100X) (#78440, Thermo Scientific, Massachusetts, US), and protein concentrations were quantified by BCA assay (Thermo Scientific, Massachusetts, US). Placental protein cytokine and chemokine levels were assessed using the Bio-Plex Pro Human Cytokine 17-plex Assay (#M5000031YV; Bio-Plex 200 system, Bio-Rad, Mississauga, Canada) according to manufacturer instructions. All protein samples were normalised to the same concentration using sample diluent. Standards with an observed/expected value outside the 80-120% range were removed, and sample duplicates with coefficients of variation >15% were excluded, per manufacturer’s protocol. Analytes with values above or below the standard curve range were excluded from analyses, resulting in 10 cytokines/chemokines for analyses: IL-1β, IL-6, IL-7, IL-8, IL-13, G-CSF, IFN-γ, MCP-1, MIP-1β, and TNF-α (expressed as pg/mL protein).

#### Placental inflammation summary score

An inflammation summary score was calculated for the pro-inflammatory placental analytes^31^ (IL-1β, IL-6, IL-7, IL-8, G-CSF, IFN-γ, MCP-1, MIP-1β, and TNF-α, excluding typically anti-inflammatory IL-13). Only samples with data available for all nine analytes were included in the summary score, for a total of 63 samples (n=41 preterm and n=22 term). For each analyte, samples were assigned a rank value ranging from one (assigned to the lowest protein expression level for that analyte) to 63 (the highest expression level, e.g. higher inflammation). Where samples had identical expression values for any given analyte, they were assigned the same rank. For each participant, the individual rank values across all nine analytes were summed to derive an overall inflammation summary score, where a greater score indicates increased levels of placental inflammation.

### Pro-inflammatory exposures/health states

To assess whether suboptimal maternal BMI with high placental inflammation associates with adverse pregnancy outcomes, binary exposure variables were derived. High placental inflammation was characterised as an inflammation summary score in the highest tertile (n=21). Due to a low number of participants with obesity and high placental inflammation, maternal overweight and obesity were considered together (Figure 1). Similarly, a combined exposure variable was derived for pregnancies with chorioamnionitis and high placental inflammation, compared to chorioamnionitis with lower placental inflammation (Figure 1).

**Figure 1.**
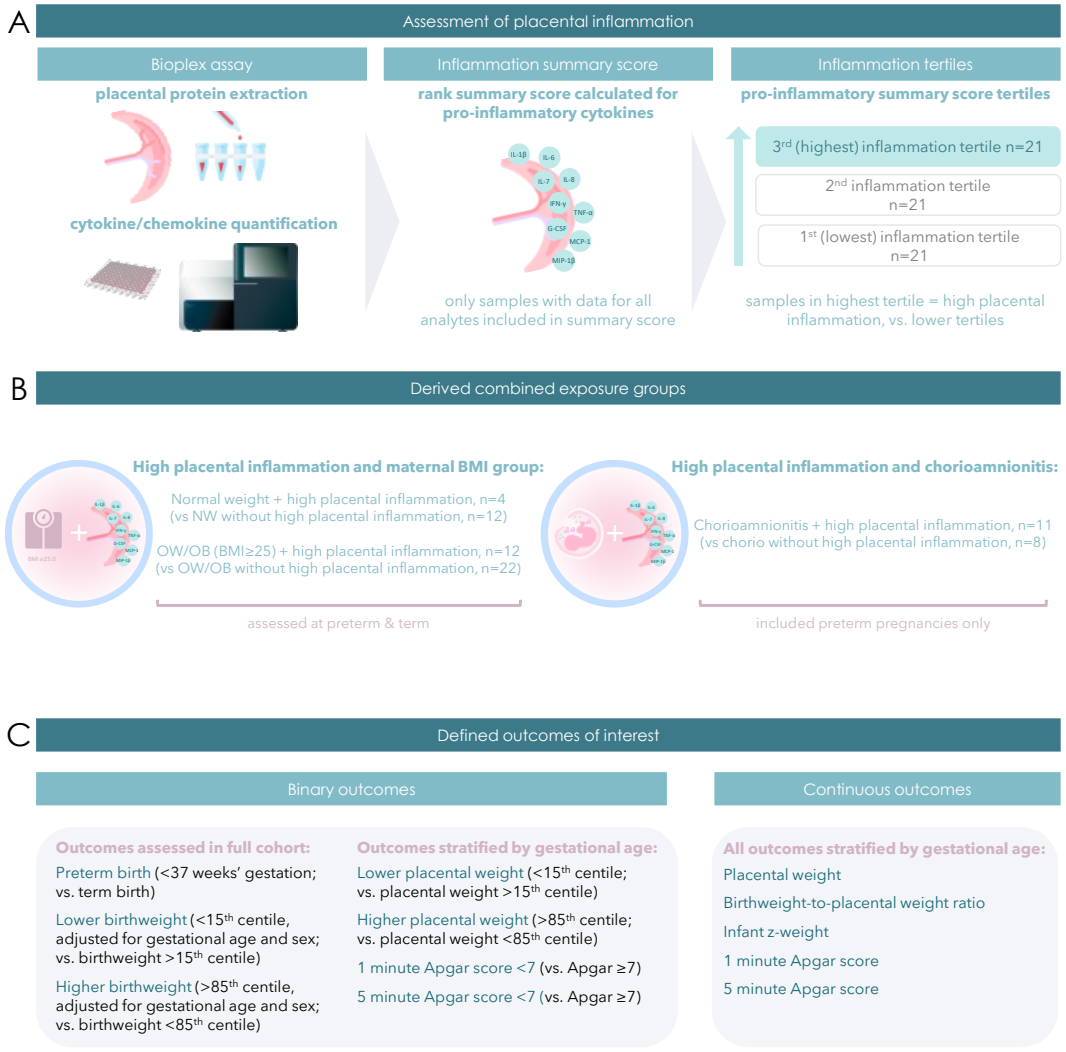
Flow diagram of methods used to measure placental inflammation, derived exposure variables, and pregnancy outcomes. **A**. Placental cytokine and chemokine protein expression levels were assessed by Bio-Plex immunoassay, and a pro-inflammatory summary score was derived. Placental samples with pro-inflammatory summary scores in the highest tertile were defined as having high placental inflammation. **B**. Combined exposure variables were derived for pregnancies with normal weight maternal BMI, and overweight/obese BMI, with high placental inflammation (highest inflammation tertile), compared to the same BMI group without high placental inflammation (lower two inflammation tertiles). Similarly, a combined exposure variable was derived for pregnancies with chorioamnionitis and high placental inflammation. All pregnancies with chorioamnionitis delivered preterm. **C**. Definitions of pregnancy, placental, and infant outcomes of interest.

### Pregnancy outcomes

Primary outcomes were PTB (<37 weeks’ gestation; binary), and placental and newborn measures at birth. The following outcomes were assessed as continuous variables: placental weight, birthweight-to-placental weight ratio (a proxy for placental efficiency), z-birthweight, and infant APGAR scores at one-and five-minutes. Binary outcome variables were also derived for low (below the 15^th^ centile) and high (above the 85^th^ centile) placental and infant weights^28,29^, and low infant Apgar scores (below seven). Infant birthweight centiles were determined based on sex and gestational age^30^. Internal placental weight centiles were derived relative to the cohort.

### Statistical analyses

#### Univariate analyses

First, we characterised placental inflammation across maternal BMI and gestational age categories, and by fetal sex. Outcomes were tested for normality and unequal variances (Levene test), and where possible, non-normal inflammatory biomarker data were log transformed to achieve normality. Data were assessed by ANOVA with Tukey *post hoc* or Welch with Games-Howell *post hoc* for normally distributed data, or Kruskal– Wallis test with Steel–Dwass *post hoc* for nonparametric data. Data were analysed using JMP statistical software (Student Edition).

### Multivariable analyses

#### Placental inflammation and pregnancy outcomes

The effects of placental inflammation, inclusive of BMI and chorioamnionitis groups, on pregnancy outcomes were assessed. Placental and infant growth and development outcomes dependent on gestational age (placental weight, birthweight-to-placental weight ratio, z-birthweight, APGAR scores) were stratified by preterm and term. Nominal logistic regression models were used to determine the associations between the placental inflammation summary score (continuous) and binary pregnancy outcome variables. Data are presented as unadjusted (model 1) (OR) or adjusted (model 2, model 3) units odds ratio (aOR) (95% CI) for odds of the adverse outcome and p-value from Effect Likelihood Ratio Test. Standard Least Squares regression models were used for continuous outcome variables. Data are reported as unadjusted (β) or adjusted parameter estimate (aβ) (95% CI) (for placental inflammation) with p-value (for the parameter estimate). Covariables in the minimally adjusted model (model 2) included infant sex (male/female), maternal age (continuous), and maternal BMI (continuous). Adjusted odds ratios and parameter estimates presented in text are from model 2. A third model, model 3, additionally adjusted for gestational age (continuous), z-gestational weight gain (continuous), antibiotic use (yes/no), chorioamnionitis status (yes/no), and glucocorticoids (yes/no).

#### Pro-inflammatory exposures

First, to address the primary objective, we assessed whether high placental inflammation associated with more adverse pregnancy outcomes across maternal BMI groups, using unadjusted and adjusted Nominal Logistic or Standard Least Squares regression models, adjusting for covariables as described above, excluding maternal BMI. Data are presented as unadjusted or adjusted odds ratio (95% CI) for odds of the adverse outcome and p-value (for BMI group + high placental inflammation) from Effect Likelihood Ratio Test, or unadjusted or adjusted parameter estimate (95% CI) for BMI group + high inflammation with p-value (for the parameter estimate). Next, to assess whether high placental inflammation contributed to adverse outcomes in pregnancies with chorioamnionitis, we used unadjusted and adjusted models as described above, with chorioamnionitis and high placental inflammation as the binary exposure group, adjusting for all covariables except for chorioamnionitis status. There were no cases of chorioamnionitis at term, thus analyses considered preterm placentae only.

## Results

### Placental cytokine/chemokine levels increased with maternal overweight and at preterm with and without chorioamnionitis

Among preterm placentae, maternal overweight associated with higher IL-6 protein expression levels compared to normal weight (p=0.02), higher placental G-CSF (p=0.02) and MCP-1 (p=0.02) levels compared to the obese group, and higher MIP-1β levels compared to the obese and underweight groups (p=0.01) (Figure 2A, Supplementary Table 1). Among term placentae, G-CSF protein expression levels were higher in overweight compared to mothers with obesity, but not different from normal weight (p=0.01; Figure 2A, Supplementary Table 1). There was no difference in the overall inflammation summary score by maternal BMI group at preterm or term (Figure 2D,E).

**Figure 2.**
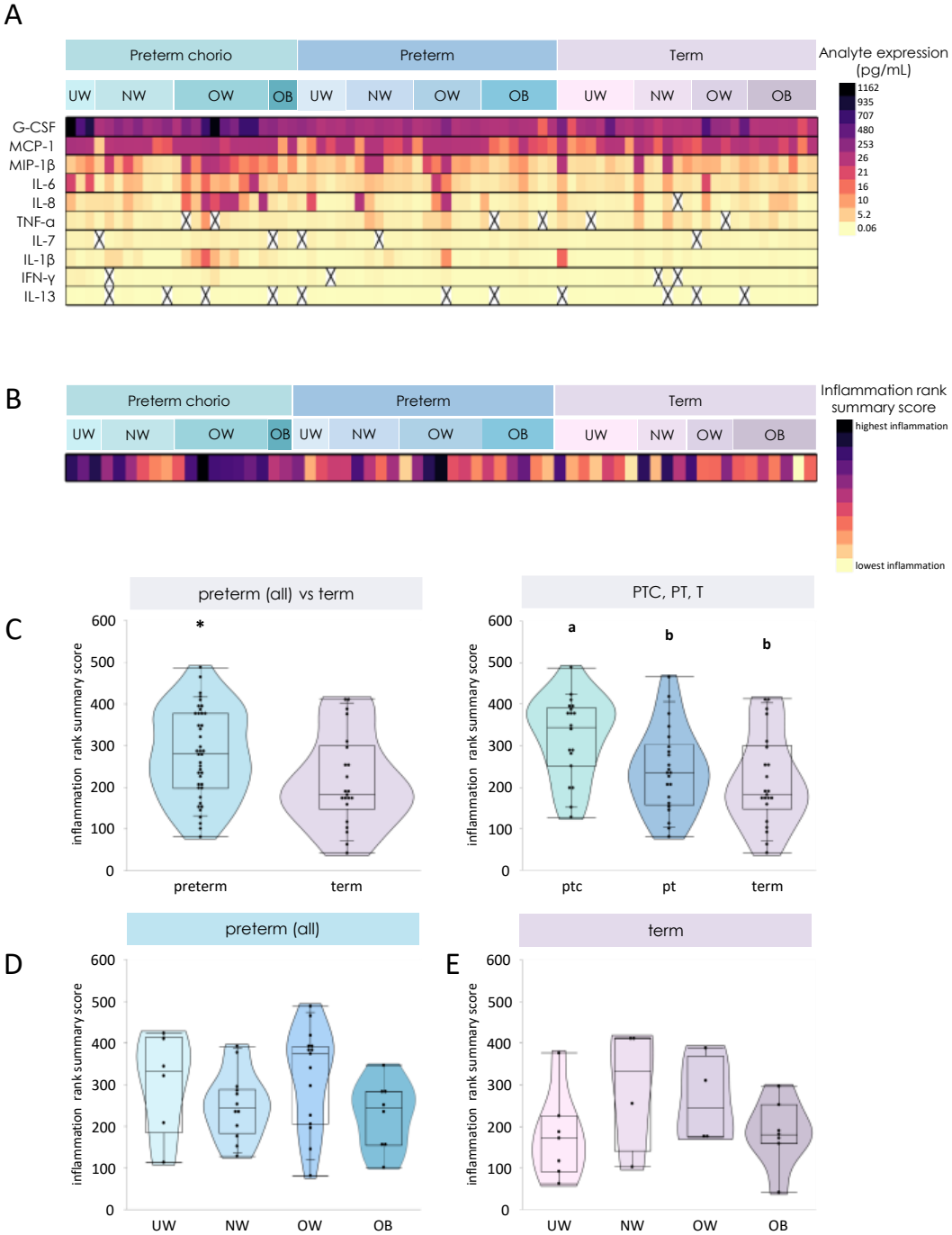
Placental cytokine and chemokine protein expression levels. **A**. Placental protein expression of IL-1β (p=0.0004), IL-6 (p=0.009), IL-7 (p=0.01), IL-13 (p=0.03), G-CSF (p=0.006), IFN-γ (p=0.046), and MIP-1β (p=0.007) were increased in all preterm placentae (with and without chorioamnionitis) compared to term placentae. When further stratifying preterm samples by preterm placentae with chorioamnionitis (PTC) and preterm placentae without chorioamnionitis (PT), IL-1β (p=0.0006), IL-6 (p=0.02), IL-8 (p<0.0001), G-CSF (p<0.0001), IFN-γ (p=0.003), MCP-1 (p=0.001), and MIP-1β (p=0.002) protein expression was increased in preterm placentae with chorioamnionitis compared to preterm without chorioamnionitis and term placentae. Placental IL-7 was also increased in preterm with chorioamnionitis compared to term (p=0.009), but not preterm without chorioamnionitis. Among preterm placentae, IL-6 protein expression levels were increased in placentae from mothers who were overweight prepregnancy, compared to normal weight (p=0.02). Placentae from mothers classified as overweight also had increased G-CSF (p=0.02) and MCP-1 (p=0.02) protein expression compared to placentae from mothers with obesity, and increased MIP-1β (p=0.01) compared to obese and underweight BMI groups, but were not different from normal weight. Among term placentae, G-CSF protein expression levels were increased in overweight compared to placentae from mothers with obesity, but not different from normal weight (p=0.01). **B, C**. The inflammation rank summary score was increased in all preterm compared to term placentae (All Preterm: 278 ± 107, Term: 216 ± 110, p=0.03), with highest inflammation among the PTC compared to preterm without chorioamnionitis and term placentae (PTC: 321 ± 98.8^A^, PT: 241 ± 102^B^, T: 216 ± 110^B^, p=0.006). **D**. There were no differences in the overall inflammation rank summary score by maternal BMI group at preterm or **E**. at term. UW = underweight. NW = normal weight. OW = overweight. OB = obese. PTC = preterm with chorioamnionitis. PT = preterm without chorioamnionitis. T = term.

Inclusive of maternal BMI, placental IL-1β (p=0.0004), IL-6 (p=0.009), IL-7 (p=0.01), IL-13 (p=0.03), G-CSF (p=0.006), IFN-γ (p=0.046), and MIP-1β (p=0.007) protein expression levels were increased in all preterm (with and without chorioamnionitis) compared to term placentae (Figure 2A, Supplementary Table 2). Further, except for IL-13, differences in cytokine levels were driven by the chorioamnionitis group (Figure 2A, Supplementary Table 3). Similarly, the overall inflammation summary score was increased in all preterm compared to term placentae (p=0.03), but highest in the preterm with chorioamnionitis subgroup (p=0.006, Figure 2B,C).

Additionally, at preterm, male placentae had decreased MCP-1 expression compared to female placentae (p=0.02, Table 1). In contrast, at term, male placentae had increased IL-1β (p=0.04) and G-CSF (p=0.02) expression compared to females (Table 1). There were no sex differences in the inflammation summary score or infant outcomes at preterm or term (Supplementary Table 4), including when further stratifying within maternal BMI and gestational age groups (Supplementary Table 5, 6).

**Table 1.** Effect of sex on protein expression levels of placental cytokines/chemokines in preterm and term pregnancies.

| Placental analyte expression | Placental sex |  |  |
|---|---|---|---|
|  | Female | Male | **p-value** |
| Preterm pregnancies | n=22 | n=29 |  |
| **IL-1β** | 0.41 (0.27, 1.68) | 0.68 (0.39, 1.21) | 0.70 |
| **IL-6** | 3.61 (2.22, 6.82) | 4.08 (2.74, 6.14) | 0.80 |
| **IL-7** | 0.98 ± 0.30 | 1.13 ± 0.41 | 0.17 |
| **IL-8** | 1.91 (0.88, 7.08) | 1.88 (1.00, 3.03) | 0.47 |
| **IL-13** | 0.15 (0.10, 0.18) | 0.15 (0.10, 0.23) | 0.54 |
| **G-CSF** | 127 (58.3, 207) | 106 (55.0, 206) | 0.43 |
| **IFN-γ** | 0.46 (0.32, 0.76) | 0.39 (0.29, 0.43) | 0.08 |
| **MCP-1** | 34.8 (22.7, 44.1) | 27.9 (14.8, 36.7) | **0.02** |
| **MIP-1β** | 8.97 (3.80, 21.7) | 8.26 (5.12, 12.2) | 0.35 |
| **TNF-α** | 1.74 (1.20, 4.00) | 1.51 (1.24, 1.95) | 0.26 |
| Term pregnancies | n=15 | n=12 |  |
| **IL-1β** | 0.28 (0.24, 0.33) | 0.33 (0.28, 0.96) | **0.04** |
| **IL-6** | 2.61 (2.06, 3.49) | 3.56 (2.56, 6.90) | 0.10 |
| **IL-7** | 0.78 ± 0.22 | 0.86 ± 0.23 | 0.41 |
| **IL-8** | 1.35 (0.77, 5.33) | 1.56 (0.78, 3.18) | 0.94 |
| **IL-13** | 0.13 (0.07, 0.18) | 0.10 (0.09, 0.15) | 0.58 |
| **G-CSF** | 41.6 (28.5, 54.7) | 77.1 (50.3, 317) | **0.02** |
| **IFN-γ** | 0.32 (0.23, 0.42) | 0.38 (0.26, 0.73) | 0.14 |
| **MCP-1** | 24.1 (16.7, 29.9) | 26.1 (20.6, 47.3) | 0.26 |
| **MIP-1β** | 4.32 (3.67, 7.70) | 5.18 (3.40, 31.5) | 0.41 |
| **TNF-α** | 1.49 (0.83, 2.44) | 2.12 (0.96, 3.69) | 0.29 |

Data are mean ± SD (ANOVA for normal distribution/equal variance data, or Welch for normal distribution/unequal variance) or median (IQR; Kruskal-Wallis/Wilcoxon test for non-parametric data). \*p<0.05.

### Placental inflammation associated with increased odds of PTB and adverse infant outcomes

We first assessed whether placental inflammation associated with impaired outcomes, inclusive of exposure groups. Higher levels of placental inflammation associated with slight increased odds of PTB (unadjusted OR: 1.01 [1.00, 1.01], p=0.03; aOR: 1.01 [1.00, 1.01], p=0.02), and slight decreased odds of higher infant birthweight (unadjusted OR: 0.99 [0.98, 1.00], p=0.02; aOR: 0.98 [0.96, 1.00], p=0.01) among the whole cohort. At preterm only, we observed slight increased odds of one-minute (unadjusted OR: 1.01 [1.00, 1.01], p=0.009; aOR: 1.01 [1.00, 1.02], p=0.004) and five-minute (OR: 1.01 [1.00, 1.02], p=0.007; aOR: 1.01 [1.00, 1.03], p=0.007) Apgar scores below seven. Placental inflammation also associated with decreased birthweight-to-placental weight ratio and infant Apgar scores as continuous outcome variables at preterm (Table 2). There were no differences in model 3, or adjusted estimates could not be reliably derived. At term, higher placental inflammation associated with decreased birthweight-to-placental weight ratio in model 3 only (Table 2).

**Table 2.** Effect of placental inflammation (continuous) on newborn and placental anthropometry and development measures at preterm and term.

|  | Unadjusted Model 1 |  | Adjusted Model 2 |  | Adjusted Model 3 |  |
|---|---|---|---|---|---|---|
|  | β (95% CI) | **p-value** | β (95% CI) | **p-value** | β (95% CI) | **p-value** |
| Preterm |  |  |  |  |  |  |
| Apgar score (1 min) | -0.009 (-0.02, -0.003) | **0.006** | -0.010 (-0.02, -0.004) | **0.004** | -0.005 (-0.01, 0.003) | 0.19 |
| Apgar score (5 min) | -0.007 (-0.01, -0.001) | **0.02** | -0.007 (-0.01, -0.001) | **0.02** | -0.002 (-0.009, 0.005) | 0.60 |
| Infant z-weight | -0.003 (-0.005, 0.00006) | 0.05 | -0.002 (-0.005, 0.0002) | 0.07 | -0.003 (-0.006, 0.001) | 0.16 |
| Placental weight (g) | -0.46 (-0.97, 0.05) | 0.08 | -0.46 (-0.97, 0.05) | 0.08 | 0.11 (-0.43, 0.66) | 0.67 |
| bw:pw | -0.005 (-0.009, -0.002) | **0.002** | -0.005 (-0.009, -0.002) | **0.003** | -0.003 (-0.007, 0.001) | 0.20 |
| Term |  |  |  |  |  |  |
| Apgar score (1 min) | 0.002 (-0.003, 0.008) | 0.39 | 0.003 (-0.003, 0.008) | 0.32 | -0.003 (-0.01, 0.004) | 0.35 |
| Apgar score (5 min) | 0.0003 (-0.001, 0.002) | 0.67 | 0.0004 (-0.0005, 0.001) | 0.37 | -0.00005 (-0.002, 0.001) | 0.94 |
| Infant z-weight | 0.00001 (-0.005, 0.005) | 1.00 | 0.0005 (-0.005, 0.006) | 0.83 | 0.003 (-0.003, 0.009) | 0.33 |
| Placental weight (g) | 0.28 (-0.75, 1.31) | 0.58 | 0.38 (-0.58, 1.34) | 0.42 | 1.18 (-0.28, 2.63) | 0.10 |
| bw:pw | -0.002 (-0.007, 0.002) | 0.32 | -0.003 (-0.007, 0.002) | 0.19 | -0.006 (-0.01, -0.00005) | **0.049** |

Data are β (95% CI) with p-value from Standard Least Squares models. Model 2 adjusted for maternal BMI, maternal age, and infant sex. Model 3 additionally adjusted for gestational age, z-gestational weight gain, antibiotic use, chorioamnionitis status, and glucocorticoid use. \*p<0.05. bw:pw = birthweight-to-placental weight ratio.

### Pregnancies with high maternal BMI and high placental inflammation had poorer outcomes

At preterm only, maternal overweight/obesity combined with high placental inflammation associated with decreased placental weight (β: -169 [-329, -9.13], p=0.04, Table 3) in model 1 only. In model 1 and model 2, maternal overweight/obesity with high placental inflammation associated with decreased placental efficiency (aβ: -1.42 [-2.29, -0.54], p=0.003), and infants with lower one-(aβ: -2.20 [-4.11, -0.29], p=0.03) and five-minute (aβ: -1.66 [-2.89, -0.42], p=0.01) Apgar scores, compared to preterm pregnancies with maternal overweight/obesity without high placental inflammation (Table 3). There were no differences at term, or in model 3. There were also no differences in infant or placental outcomes in pregnancies with normal BMI and high placental inflammation compared to normal BMI without high placental inflammation at preterm or term (Table 3).

**Table 3.** Effect of maternal prepregnancy BMI combined with high placental inflammation on newborn and placental anthropometry and development measures at preterm and term.

|  | NW + high placental inflammation |  |  |  |  |  | OW/OB + high placental inflammation |  |  |  |  |  |
|---|---|---|---|---|---|---|---|---|---|---|---|---|
|  | Model 1<br>Unadjusted β<br>(95% CI) | p-value | Model 2<br>Adjusted β<br>(95% CI) | p-value | Model 3<br>Adjusted β<br>(95% CI) | p-value | Model 1<br>Unadjusted β<br>(95% CI) | p-value | Model 2<br>Adjusted β<br>(95% CI) | p-value | Model 3<br>Adjusted β<br>(95% CI) | p-value |
| Preterm |  |  |  |  |  |  |  |  |  |  |  |  |
| Apgar score (1 min) | -0.30 (-4.80, 4.20) | 0.89 | -0.78 (-6.14, 4.57) | 0.74 | 0.42 (-9.12, 9.96) | 0.90 | -2.38 (-4.18, -0.58) | **0.01** | -2.20 (-4.11, -0.29) | **0.03** | -3.90 (-8.44, 0.63) | 0.08 |
| Apgar score (5 min) | -2.5 (-8.19, 3.19) | 0.35 | -2.26 (-8.31, 3.80) | 0.42 | -0.78 (-7.92, 6.36) | 0.75 | -1.65 (-2.78, -0.51) | **0.007** | -1.66 (-2.89, -0.42) | **0.01** | -0.70 (-3.40, 2.01) | 0.57 |
| Infant z-weight | -0.36 (-1.83, 1.10) | 0.59 | -0.32 (-2.10, 1.47) | 0.69 | 1.00 (-2.68, 4.69) | 0.45 | -0.55 (-1.26, 0.16) | 0.12 | -0.58 (-1.34, 0.18) | 0.13 | 0.02 (-1.60, 1.65) | 0.97 |
| Placental weight (g) | 38.0 (-198, 274) | 0.73 | 8.60 (-265, 282) | 0.94 | 125 (-357, 607) | 0.47 | -169 (-329, -9.13) | **0.04** | -164 (-338, 10.2) | 0.06 | -137 (-563, 288) | 0.48 |
| bw:pw | -0.46 (-2.89, 1.97) | 0.68 | -0.25 (-3.35, 2.86) | 0.86 | -1.53 (-7.35, 4.29) | 0.46 | -1.43 (-2.24, -0.62) | **0.002** | -1.42 (-2.29, -0.54) | **0.003** | 0.70 (-2.20, 3.59) | 0.59 |
| Term |  |  |  |  |  |  |  |  |  |  |  |  |
| Apgar score (1 min) | 0.50 (-1.65, 2.65) | 0.42 | -1.3 (-9.44, 6.84) | 0.29 | - |  | 1.25 (-2.00, 4.50) | 0.40 | 1.61 (-2.50, 5.71) | 0.38 | -1.12 (-16.3, 14.1) | 0.78 |
| Apgar score (5 min) | 0 | - | 0 | - | - |  | 0.25 (-0.54, 1.04) | 0.49 | 0.42 (-0.44, 1.29) | 0.27 | -0.34 (-3.84, 3.16) | 0.72 |
| Infant z-weight | 0.98 (-0.12, 2.08) | 0.06 | 1.89 (-2.50, 6.29) | 0.12 | - |  | -1.36 (-4.17, 1.45) | 0.30 | -1.32 (-4.95, 2.31) | 0.42 | 1.62 (-5.64, 8.88) | 0.53 |
| Placental weight (g) | 155 (-3036, 3346) | 0.65 | - |  | - |  | -106 (-597, 385) | 0.64 | -63.7 (-688, 560) | 0.82 | 477 (-1262, 2216) | 0.45 |
| bw:pw | -1.14 (-29.1, 26.8) | 0.69 | - |  | - |  | 0.08 (-1.52, 1.69) | 0.91 | 0.008 (-2.05, 2.06) | 0.99 | -0.96 (-3.95, 2.03) | 0.38 |

Data are β (95% CI) for NW pregnancies with high placental inflammation, and OW/OB pregnancies with high placental inflammation, respectively, and p-value from Standard Least Squares models. Model 2 adjusted for maternal age and infant sex. Model 3 additionally adjusted for gestational age, z-gestational weight gain, antibiotic use, chorioamnionitis status, and glucocorticoid use. \*p<0.05. Among term normal weight pregnancies, there was no variability in 5-minute Apgar scores across pregnancies with and without high placental inflammation. Estimates could not be reliably derived for placental weight or bw:pw in Model 2, or any outcomes in Model 3 for term normal weight pregnancies. NW = normal weight. OW/OB = overweight/obesity. bw:pw = birthweight-to-placental weight ratio.

### Pregnancies with chorioamnionitis and high placental inflammation did not have altered outcomes

Pregnancies complicated by chorioamnionitis combined with high placental inflammation did not have poorer infant or placental outcomes compared to pregnancies with chorioamnionitis and lower placental inflammation (Table 4). As no term pregnancies were diagnosed with chorioamnionitis in this cohort, odds of PTB could not be assessed.

**Table 4.**
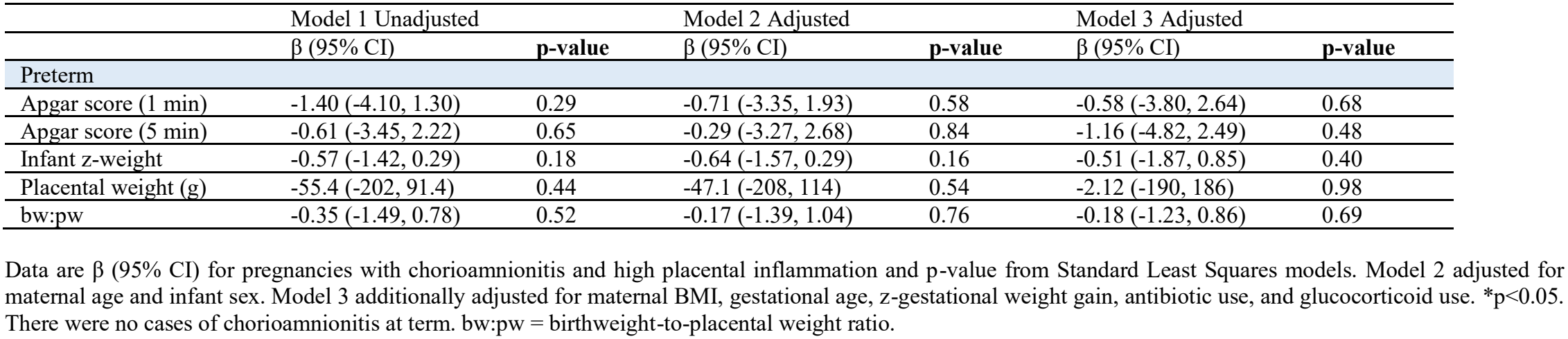
Effect of chorioamnionitis combined with high placental inflammation on newborn and placental anthropometry and development measures at preterm.

## Discussion

### Principal Findings

Among preterm pregnancies without other major maternal comorbidities, maternal overweight/obesity with high placental inflammation was associated with lower placental efficiency and lower infant Apgar scores, compared with maternal overweight/obesity without high placental inflammation. These associations were robust to adjustment for maternal age and infant sex, and attenuated only in a further adjusted model that included likely mediators. In contrast, high placental inflammation in pregnancies of normal BMI was not associated with adverse outcomes. Within the chorioamnionitis preterm group, high placental inflammation did not worsen adverse pregnancy outcomes beyond those already seen with chorioamnionitis and lower levels of placental inflammation. These findings suggest that pregnancies with elevated maternal BMI may have poorer outcomes when mothers have inflamed placentae, particularly at preterm.

### Results

Preterm pregnancies complicated by maternal overweight/obesity and high placental inflammation had lower placental efficiency, and newborns with decreased one-and five-minute Apgar scores. This suggests that high placental inflammation may be one factor that signals increased risk for adverse outcomes among pregnancies complicated by maternal overweight/obesity. Both high BMI and inflammation have been independently linked with less efficient placentae^38^, in line with our findings. Decreased efficiency can compromise placental resource allocation, and has also been linked with later health outcomes for the child, such as cardiometabolic risk^40^. Despite these placental alterations, no changes in infant birthweight were seen, which may suggest other placental adaptations to ensure fetal growth. Reports on the effect of BMI on Apgar scores are more inconsistent: a systematic review found that overweight and obesity associated with low infant Apgar scores^41^, similar to a cohort study that assessed high BMI among preterm pregnancies only^43^, however, both included cohorts with additional comorbidities. In contrast, another study that included only glucose tolerant, non-hypertensive women with obesity found no differences in Apgar scores, however only investigated term pregnancies^42^. Our findings showed that infants born to mothers with overweight/obesity and high placental inflammation had lower Apgar scores, at preterm ages only. As fewer placentae had high inflammation at term, additional studies would be needed to confirm findings at later gestational ages. Collectively, these results suggest that even in high maternal BMI without additional maternal metabolic comorbidities, there is some risk for adverse outcomes. Having an inflamed placenta may help to distinguish pregnancies with high BMI at greatest risk for impaired fetal development.

Contrary to our hypothesis, placentae from mothers who were overweight, not those with obesity, had increased levels of pro-inflammatory cytokines. In part, this finding could be explained by the greater number of chorioamnionitis cases in the overweight group compared to the obese group, because chorioamnionitis associated with highest levels of inflammation. Further, mothers who were overweight received fewer antibiotics, compared to mothers with obesity who received the greatest proportion of antibiotics, which may have attenuated inflammation in the obese BMI group^44^. While there were no differences in the overall inflammation summary score between BMI groups, this suggests that overweight BMI, and not just obesity, can impact placental function and lead to potential negative implications for the pregnancy.

In pregnancies with normal BMI, high placental inflammation was not associated with worse placental or neonatal outcomes. This contrasts with the findings in pregnancies affected by overweight/obesity, in which high placental inflammation was accompanied by poorer outcomes. However, these findings should be interpreted cautiously given the smaller number of normal-BMI pregnancies with high placental inflammation in our cohort. If replicated, this may suggest that uncomplicated pregnancies are less susceptible to adversity associated with high placental inflammation, though the underlying cause of inflammation may still be clinically relevant.

Pregnancies complicated by chorioamnionitis that had high placental inflammation did not have poorer outcomes compared to pregnancies with chorioamnionitis and placental cytokine output in the lowest two tertiles. Chorioamnionitis is diagnosed histologically based on placental neutrophil infiltration, and is a known contributor to PTB and impaired fetoplacental development^45^. Overall, placentae with chorioamnionitis did have increased cytokine and chemokine levels compared to preterm without chorioamnionitis and term placentae, supporting an increased inflammatory response, as expected. This is consistent with other studies showing increased inflammatory mediators in pregnancy tissues and fluids in chorioamnionitis cases, such as within amniotic fluid and umbilical serum^46-48^. It is possible that the local influx of inflammatory infiltrates^50,51^ has subsequent impacts on pregnancy and birth outcomes, irrespective of further elevated levels of pro-inflammatory placental cytokines and chemokines. Alternatively, chorioamnionitis itself is a substantial insult, and the outcomes it produces may leave little room for additional cytokine burden to exert a measurable effect in a sample of this size. While other studies have shown mixed results on the effects of cytokines on birth outcomes in chorioamnionitis-affected pregnancies, chorioamnionitis can progress to elicit a fetal immune response (FIRS), which has been linked with poorer fetal development^52^. This suggests that inflammation involving the umbilical cord or fetal circulation may be more closely related to adverse neonatal outcomes than placental cytokine levels alone. Furthermore, the inflammatory processes associated with maternal overweight/obesity may differ from those occurring in chorioamnionitis. Chronic, low-grade metabolic inflammation in pregnancies affected by overweight/obesity may have different implications for neonatal outcomes than the acute inflammatory response associated with chorioamnionitis.

### Clinical Implications

Overweight and obesity in pregnancy and PTB remain prevalent conditions and contribute to immediate and longer-term child health challenges^53,54^, therefore, identifying subgroups at greatest risk for poor outcomes may inform targeted surveillance and new interventions. BMI alone has limited predictive value for identifying pregnancies at risk of additional complications^22^, while much of the available evidence comes from cohorts with coexisting maternal comorbidities. Our findings raise the possibility that an inflamed placental phenotype marks a higher-risk subgroup within pregnancies affected by overweight/obesity, and future research should address whether this is a clinically useful stratifying factor.

### Research Implications

Our study reveals three research priorities. First, the associations we tested need replication in larger cohorts and with overweight and obesity assessed separately. Second, because placental inflammation is measured at delivery, future studies should investigate whether inflammatory markers in placental biopsies collected earlier in pregnancy, or circulating maternal or fetal inflammatory biomarkers that can be assessed antenatally, can identify pregnancies at risk of adverse outcomes. Third, determining factors that distinguish pregnancies with and without high placental inflammation, such as medication exposures, and placental sex^55^, would clarify what drives the phenotype.

### Strengths and Limitations

Strengths include a cohort selected to exclude other major maternal comorbidities, among both preterm and term deliveries, and assessment of a panel of cytokines and chemokines rather than histopathology or a single inflammatory marker. The main limitation is statistical power. Our exposure of interest included pregnancies with the highest levels of placental inflammation, further stratified by BMI or chorioamnionitis groups, resulting in fewer participants per exposure group. This smaller sample size also meant the fully adjusted analysis (model 3) likely had more covariables than size could support, leading to less precise, or unstable estimates. However, most associations in model 1 retained statistical significance in the minimally adjusted model 2, supporting these findings, while the direction of associations were generally similar in model 3. Analyses were also limited by fewer normal weight BMI, and term pregnancies exhibiting high placental inflammation, and sample size was insufficient to explore maternal underweight with high inflammation.

### Conclusions

In pregnancies complicated by overweight or obesity, but without other major maternal comorbidities, a high placental inflammatory profile may help identify those at greatest risk of adverse birth outcomes at preterm. Testing this in larger cohorts will help determine whether placental inflammation, or an antenatal correlate of this phenotype, could improve risk stratification beyond BMI alone and inform better surveillance, and early, individualised prenatal care.

## Data availability

Some or all datasets generated during and/or analysed during the current study are not publicly available but are available from the corresponding author on reasonable request.

## Author Contributions

Conceptualization, methodology: KLC, HS, EB; investigation and formal analysis: HS; writing—original draft preparation: HS, KLC; writing—review and editing: HS, KLC, EB; funding acquisition, resources, supervision: KLC.

## Acknowledgments

We thank the donors, the RCWIH BioBank, the Lunenfeld-Tanenbaum Research Institute and the Mount Sinai Hospital/UHN Department of Obstetrics and Gynaecology for the human specimens used in this study (https://biobank.lunenfeld.ca).

## Supplementary tables

**Supplementary Table 1.** Effect of maternal prepregnancy BMI on protein expression levels of placental cytokines/chemokines in preterm and term pregnancies.

| Placental analyte expression | Prepregnancy BMI |  |  |  |  |
|---|---|---|---|---|---|
|  | UW | NW | OW | OB | p-value |
| Preterm pregnancies | n=8 | n=15 | n=17 | n=11 |  |
| **IL-1β** | 0.41 (0.24, 1.18)<br>n=8 | 0.81 (0.4, 2.06)<br>n=15 | 1.31 (0.40, 5.01)<br>n=16 | 0.39 (0.28, 1.07)<br>n=11 | 0.10 |
| **IL-6** | 3.54 (2.69, 16.1)<sup>AB</sup><br>n=8 | 4.06 (1.86, 6.00)<sup>A</sup><br>n=15 | 6.40 (3.65, 13.3)<sup>B</sup><br>n=16 | 3.21 (2.33, 4.67)<sup>AB</sup><br>n=11 | **0.02** |
| **IL-7** | 1.17 ± 0.37<br>n=7 | 0.88 ± 0.19<br>n=12 | 1.12 ± 0.38<br>n=17 | 1.09 ± 0.44<br>n=10 | 0.27 |
| **IL-8** | 2.12 (0.61, 3.16)<br>n=7 | 1.88 (1.28, 3.45)<br>n=14 | 7.99 (1.11, 26.0)<br>n=16 | 1.21 (0.84, 2.94)<br>n=10 | 0.10 |
| **IL-13** | 0.16 (0.10, 0.24)<br>n=7 | 0.14 (0.10, 0.17)<br>n=12 | 0.15 (0.10, 0.18)<br>n=15 | 0.15 (0.11, 0.24)<br>n=9 | 0.92 |
| **G-CSF** | 175 (61.8, 805)<sup>AB</sup><br>n=8 | 140 (77.5, 182)<sup>AB</sup><br>n=15 | 207 (84.8, 587)<sup>A</sup><br>n=17 | 63.6 (41.9, 136)<sup>B</sup><br>n=11 | **0.02** |
| **IFN-γ** | 0.84 (0.34, 1.29)<br>n=7 | 0.39 (0.25, 0.42)<br>n=13 | 0.43 (0.32, 0.50)<br>n=14 | 0.34 (0.27, 0.56)<br>n=11 | 0.06 |
| **MCP-1** | 31.5 (14.6, 73.6)<sup>AB</sup><br>n=8 | 27.9 (18.5, 36.8)<sup>AB</sup><br>n=15 | 36.7 (27.6, 39.4)<sup>A</sup><br>n=15 | 18.5 (14.9, 33.1)<sup>B</sup><br>n=11 | **0.02** |
| **MIP-1β** | 6.43 (4.70, 8.64)<sup>A</sup><br>n=8 | 8.08 (3.76, 19.4)<sup>AB</sup><br>n=15 | 13.4 (9.61, 22.4)<sup>B</sup><br>n=16 | 5.18 (3.84, 11.3)<sup>A</sup><br>n=11 | **0.01** |
| **TNF-α** | 1.43 (0.90, 1.60)<br>n=7 | 1.61 (1.35, 3.84)<br>n=15 | 1.63 (1.19, 3.09)<br>n=14 | 1.95 (1.23, 3.05)<br>n=9 | 0.39 |
| Term pregnancies | n=8 | n=6 | n=6 | n=7 |  |
| **IL-1β** | 0.28 (0.26, 0.33)<br>n=7 | 0.63 (0.26, 1.28)<br>n=6 | 0.28 (0.22, 0.54)<br>n=6 | 0.34 (0.31, 0.36)<br>n=7 | 0.32 |
| **IL-6** | 2.70 (1.72, 3.64)<br>n=7 | 3.08 (1.97, 8.67)<br>n=6 | 2.52 (2.27, 4.20)<br>n=5 | 3.41 (2.13, 3.88)<br>n=7 | 0.69 |
| **IL-7** | 0.73 ± 0.13<br>n=8 | 0.90 ± 0.27<br>n=6 | 0.94 ± 0.31<br>n=5 | 0.86 ± 0.30<br>n=7 | 0.47 |
| **IL-8** | 1.35 (0.85, 1.57)<br>n=7 | 1.56 (0.67, 6.86)<br>n=5 | 1.51 (0.66, 4.89)<br>n=6 | 1.77 (0.78, 6.56)<br>n=7 | 0.67 |
| **IL-13** | 0.11 (0.09, 0.14)<br>n=7 | 0.10 (0.08, 0.24)<br>n=5 | 0.10 (0.08, 0.19)<br>n=4 | 0.14 (0.10, 0.18)<br>n=7 | 0.82 |
| **G-CSF** | 84.5 (43.6, 301)<sup>AB</sup><br>n=8 | 54.5 (36.7, 85.3)<sup>AB</sup><br>n=6 | 197 (59.4, 497)<sup>A</sup><br>n=6 | 37.8 (27.3, 42.7)<sup>B</sup><br>n=7 | **0.01** |
| **IFN-γ** | 0.27 (0.24, 0.38)<br>n=8 | 0.54 (0.32, 0.88)<br>n=4 | 0.32 (0.28, 0.61)<br>n=6 | 0.40 (0.21, 0.44)<br>n=6 | 0.15 |
| **MCP-1** | 19.0 (11.1, 29.8)<br>n=8 | 23.4 (19.4, 56.3)<br>n=6 | 31.0 (20.7, 38.1)<br>n=6 | 25.7 (19.2, 27.9)<br>n=6 | 0.22 |
| **MIP-1β** | 3.67 (2.85, 3.80)<br>n=7 | 7.83 (3.78, 50.8)<br>n=6 | 7.42 (4.02, 7.96)<br>n=6 | 4.28 (3.69, 7.82)<br>n=7 | 0.08 |
| **TNF-α** | 1.48 (1.25, 2.45)<br>n=7 | 2.96 (0.88, 6.58)<br>n=6 | 0.96 (0.81, 2.56)<br>n=5 | 2.02 (0.98, 2.90)<br>n=7 | 0.43 |

Data are mean ± SD (ANOVA for normal distribution/equal variance data with Tukey post hoc, or Welch for normal distribution/unequal variance with Games-Howell post hoc) or median (IQR; Kruskal-Wallis/Wilcoxon test for non-parametric data with Steel–Dwass post hoc). Post hoc differences between groups are denoted by different letters. \*p<0.05. UW = underweight. NW = normal weight. OW = overweight. OB = obese.

**Supplementary Table 2.** Effect of gestational age on protein expression levels of placental cytokines/chemokines.

| Placental analyte expression | Gestational age group |  |  |
|---|---|---|---|
|  | Preterm<br>(n=51) | Term<br>(n=27) | **p-value** |
| **IL-1β** | 0.56 (0.31, 1.34)<br>n=45 | 0.30 (0.26, 0.35)<br>n=23 | **0.0004** |
| **IL-6** | 3.70 (2.47, 6.32)<br>n=46 | 2.80 (2.13, 3.64)<br>n=23 | **0.009** |
| **IL-7** | 1.04 ± 0.34<br>n=45 | 0.84 ± 0.25<br>n=26 | **0.01** |
| **IL-8** | 1.87 (0.92, 3.39)<br>n=43 | 1.35 (0.78, 2.86)<br>n=24 | 0.20 |
| **IL-13** | 0.15 (0.10, 0.18)<br>n=42 | 0.11 (0.08, 0.17)<br>n=22 | **0.03** |
| **G-CSF** | 111.8 (57.8, 198.2)<br>n=44 | 45.5 (37.8, 83.6)<br>n=23 | **0.006** |
| **IFN-γ** | 0.4 (0.30, 0.55)<br>n=44 | 0.32 (0.25, 0.41)<br>n=22 | **0.046** |
| **MCP-1** | 29.4 ± 13.8<br>n=48 | 24.8 ± 9.72<br>n=25 | 0.10 |
| **MIP-1β** | 7.65 (4.46, 12.0)<br>n=45 | 4.23 (3.37, 7.56)<br>n=24 | **0.007** |
| **TNF-α** | 1.60 (1.21, 2.46)<br>n=44 | 1.49 (0.92, 2.79)<br>n=24 | 0.55 |

Data are mean ± SD (ANOVA; normal distribution/equal variance or Welch; normal distribution/unequal variance) or median (IQR; Kruskal-Wallis/Wilcoxon test for non-parametric data). \*p<0.05.

**Supplementary Table 3.**
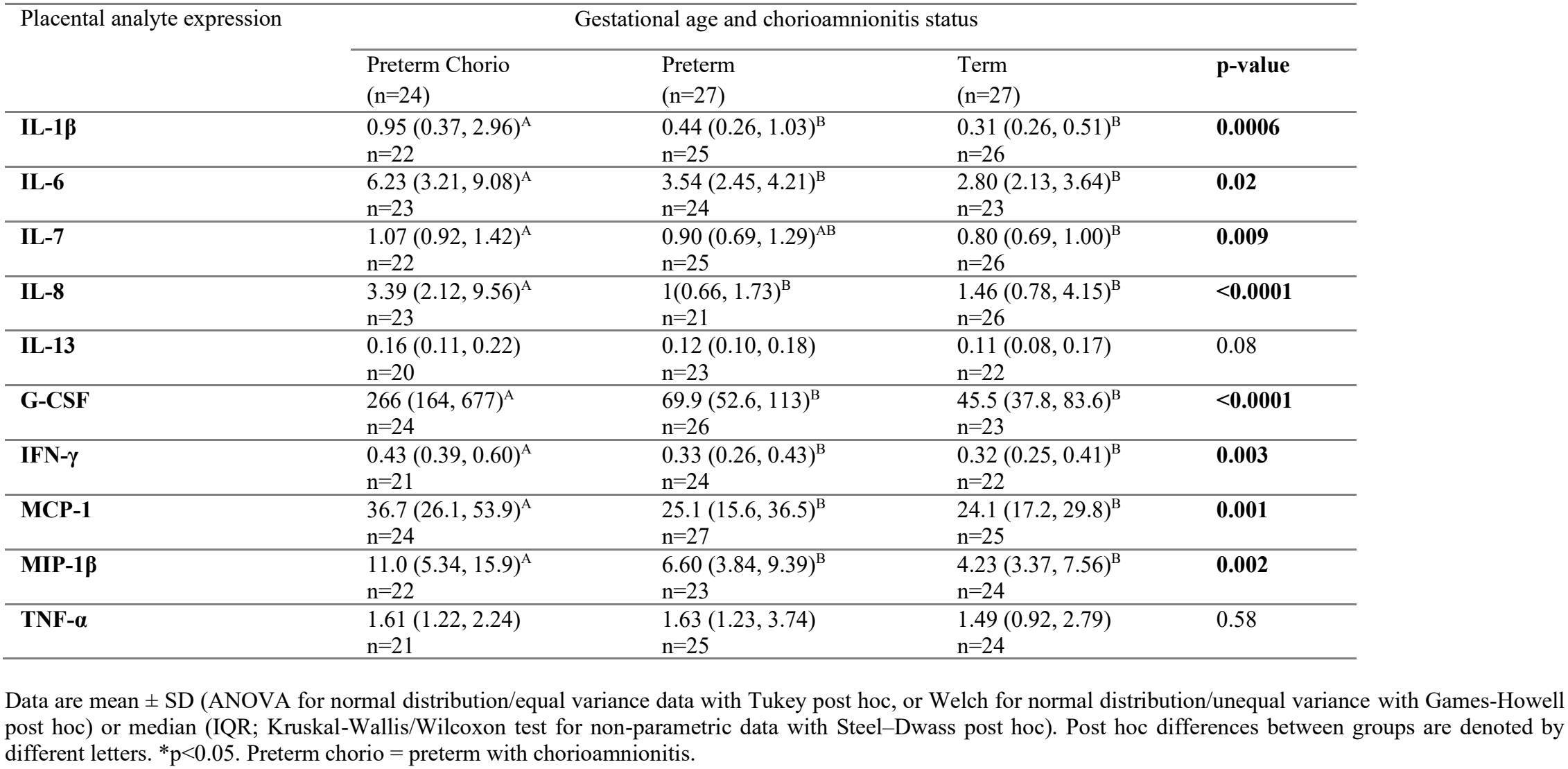
Effect of gestational age and chorioamnionitis status on protein expression levels of placental cytokines/chemokines.

**Supplementary Table 4.** Effect of sex on newborn and placental anthropometry and development measures in preterm and term pregnancies.

|  | Placental sex |  |  |
|---|---|---|---|
|  | Female | Male | **p-value** |
| Preterm pregnancies | n=17 | n=24 |  |
| **Placental inflammation** |  |  |  |
| Inflammation summary score | 286 ± 123 | 273 ± 95.8 | 0.71 |
| **Infant/placental outcomes** |  |  |  |
| Apgar score (1 min) | 8 (5.5, 9) | 8 (6, 9) | 0.85 |
| Apgar score (5 min) | 9 (8, 9) | 8.5 (8, 9) | 0.23 |
| Infant z-weight | 0.07 ± 1.18 | 0.15 ± 0.75 | 0.79 |
| Placental weight (g) | 507 ± 228 | 467 ± 128 | 0.52 |
| bw:pw | 3.97 ± 1.25 | 4.16 ± 1.19 | 0.62 |
| Term pregnancies | n=10-11 | n=11 |  |
| **Placental inflammation** |  |  |  |
| Inflammation summary score | 191 ± 97.0 | 241 ± 121 | 0.30 |
| **Infant/placental outcomes** |  |  |  |
| Apgar score (1 min) | 9 (7.5, 9) | 9 (8, 9) | 1.00 |
| Apgar score (5 min) | 9 (9, 9) | 9 (9, 9) | 0.94 |
| Infant z-weight | 0.43 (-0.09, 1.29) | -0.06 (-0.71, 0.74) | 0.20 |
| Placental weight (g) | 650 (570, 850) | 560 (458, 743) | 0.07 |
| bw:pw | 5.22 ± 0.85 | 6.01 ± 1.09 | 0.08 |

Data are mean ± SD (ANOVA for normal distribution/equal variance data, or Welch for normal distribution/unequal variance) or median (IQR; Kruskal-Wallis/Wilcoxon test for non-parametric data). \*p<0.05.

**Supplementary Table 5.** Effect of sex on newborn and placental anthropometry and development measures across maternal prepregnancy BMI groups in preterm and term pregnancies.

|  | **UW** |  |  | **NW** |  |  | **OW** |  |  | **OB** |  |  |
|---|---|---|---|---|---|---|---|---|---|---|---|---|
|  | Female | Male | p-value | Female | Male | p-value | Female | Male | p-value | Female | Male | p-value |
| Preterm pregnancies | n=2 | n=4 |  | n=4 | n=8 |  | n=8 | n=7 |  | n=3 | n=5 |  |
| **Placental inflammation** |  |  |  |  |  |  |  |  |  |  |  |  |
| Inflammation summary score | 377 ± 46.7 | 266 ± 135 | 0.34 | 223 ± 112 | 264 ± 65.9 | 0.45 | 324 ± 132 | 311 ± 117 | 0.85 | 206 ± 93.5 | 239 ± 82.9 | 0.62 |
| **Infant/placental outcomes** |  |  |  |  |  |  |  |  |  |  |  |  |
| Apgar score (1 min) | 5 (2, 8) | 8.5 (7.25, 9) | 0.23<sup>#</sup> | 8.5 (8, 9) | 7.5 (3.75, 9) | 0.47 | 7 (5, 8.75) | 8 (4, 9) | 0.68 | 9 (7, 9) | 8 (4.5, 9) | 0.43<sup>#</sup> |
| Apgar score (5 min) | 7.5 (6, 9) | 8.5 (8, 9) | 0.62<sup>#</sup> | 9 | 8.5 (2, 9) | 0.11 | 8.5 (6.5, 9) | 8 (8, 9) | 0.90 | 9 | 9 (8, 9) | 0.24<sup>#</sup> |
| Infant z-weight | -1.41 ± 1.94 | -0.23 ± 0.72 | 0.31 | 0.24 ± 0.96 | -0.18 ± 0.77 | 0.43 | 0.15 ± 0.89 | 0.33 ± 0.48 | 0.64 | 0.58 ± 1.57 | 0.71 ± 0.82 | 0.88 |
| Placental weight (g) | 288 ± 124 | 470 ± 29.4 | 0.28 | 513 ± 135 | 394 ± 118 | 0.15 | 541 ± 242 | 472 ± 125 | 0.49 | 553 ± 348 | 576 ± 143 | 0.90 |
| bw:pw | 4.12 ± 2.17 | 4.81 ± 0.47 | 0.73 | 4.06 ± 0.53 | 4.19 ± 1.66 | 0.88 | 3.62 ± 1.40 | 3.79 ± 1.22 | 0.80 | 4.70 ± 1.23 | 4.11 ± 0.51 | 0.37 |
| Term pregnancies | n=2 | n=5 |  | n=0 | n=3-4 |  | n=3 | n=1 |  | n=5-6 | n=1 |  |
| **Placental inflammation** |  |  |  |  |  |  |  |  |  |  |  |  |
| Inflammation summary score | 125 ± 87.7 | 196 ± 113 | 0.47 | - | 295 ± 148 | - | 246 ± 122 | 310 | - | 185 ± 87.3 | 180 | - |
| **Infant/placental outcomes** |  |  |  |  |  |  |  |  |  |  |  |  |
| Apgar score (1 min) | 8.5 (8, 9) | 9 (8, 9) | 0.82<sup>#</sup> | - | 9 (8.25, 9) | - | 9 | 9 | - | 7.4 ± 2.30 | 7 | - |
| Apgar score (5 min) | 9 | 9 | - | - | 9 | - | 9 | 9 | - | 9 (8.5, 9) | 8 | - |
| Infant z-weight | 0.21 ± 0.62 | -0.33 ± 0.46 | 0.25 | - | 0.26 ± 0.60 | - | 0.67 ± 0.58 | -1.14 | - | 1.06 ± 1.99 | 1.89 | - |
| Placental weight (g) | 625 ± 35.4 | 522 ± 96.0 | 0.22 | - | 647 ± 170 | - | 770 ± 144 | 520 | - | 610 (565, 1000) | 800 | - |
| bw:pw | 5.36 ± 0.32 | 6.52 ± 0.95 | 0.17 | - | 5.62 ± 1.43 | - | 5.07 ± 0.82 | 5.73 | - | 5.24 ± 1.06 | 4.93 | - |

Data are mean ± SD (ANOVA for normal distribution/equal variance data, or Welch for normal distribution/unequal variance) or median (IQR; Kruskal-Wallis/Wilcoxon test for non-parametric data). \*p<0.05. <sup>#</sup>Small sample size. UW = underweight. NW = normal weight. OW = overweight. OB = obese.

**Supplementary Table 6.** Effect of sex on newborn and placental anthropometry and development measures in preterm with chorioamnionitis, preterm without chorioamnionitis, and term pregnancies.

|  | **Preterm Chorio** |  |  | **Preterm** |  |  | **Term** |  |  |
|---|---|---|---|---|---|---|---|---|---|
|  | Female<br>n=8 | Male<br>n=11 | p-value | Female<br>n=9 | Male<br>n=13 | p-value | Female<br>n=11 | Male<br>n=10-11 | p-value |
| **Placental inflammation** |  |  |  |  |  |  |  |  |  |
| Inflammation summary score | 297 ± 130 | 338 ± 70.6 | 0.44 | 275 ± 124 | 218 ± 79.5 | 0.20 | 191 ± 97.0 | 241 ± 121 | 0.30 |
| **Infant/placental outcomes** |  |  |  |  |  |  |  |  |  |
| Apgar score (1 min) | 8 (4.5, 9) | 7 (3, 8) | 0.38 | 8 (6, 9) | 9 (7, 9) | 0.20 | 9 (7.5, 9) | 9 (8, 9) | 1.00 |
| Apgar score (5 min) | 9 (6.5, 9) | 8 (2, 9) | 0.13 | 9 (8.5, 9) | 9 (8, 9) | 0.80 | 9 (9, 9) | 9 (9, 9) | 0.95 |
| Infant z-weight | -0.20 (-0.49, 058.) | 0.12 (-0.34, 0.31) | 0.43 | 0.30 ± 1.11 | 0.17 ± 0.93 | 0.77 | 0.43 (-0.09, 1.29) | -0.06 (-0.71, 0.74) | 0.20 |
| Placental weight (g) | 358 (263, 466) | 400 (350, 470) | 0.39 | 599 ± 219 | 509 ± 136 | 0.24 | 650 (570, 850) | 560 (458, 743) | 0.07 |
| bw:pw | 3.74 ± 1.44 | 3.44 ± 0.92 | 0.58 | 4.03 (3.35, 4.49) | 4.71 (3.89, 5.21) | 0.12 | 5.22 ± 0.85 | 6.01 ± 1.09 | 0.08 |

Data are mean ± SD (ANOVA for normal distribution/equal variance data, or Welch for normal distribution/unequal variance) or median (IQR; Kruskal-Wallis/Wilcoxon test for non-parametric data). \*p<0.05. Preterm chorio = preterm with chorioamnionitis. bw:pw = birthweight-to-placental weight ratio.

